# Comparison of Object Detection Approaches Applied to Field Images of Papanicolaou Stained Cytology Slides

**DOI:** 10.1101/2021.08.25.21262605

**Authors:** André Victória Matias, Allan Cerentini, Luiz Antonio Buschetto Macarini, João Gustavo Atkinson Amorim, Felipe Perozzo Daltoé, Aldo von Wangenheim

## Abstract

Papanicolaou is an inexpensive and non-invasive method, generally applied to detect cervical cancer, that can also be useful to detect cancer on oral cavities. Although oral cancer is considered a global health issue with 350.000 people diagnosed over a year it can successfully be treated if diagnosed at early stages. The manual process of analyzing cells to detect abnormalities is time-consuming and subject to variations in perceptions from different professionals. To evaluate a possible solution to the automation of this process, in this paper we employ the object detection deep learning approach in the analysis of this type of image using 3 models: RetinaNet, Faster R-CNN, and Mask R-CNN. We trained and tested the models using images from 6 cytology slides (4 cancer cases and 2 healthy samples) and our results show that Mask R-CNN was the best model for localization and classification of nuclei with an IoU of 0.51 and recall of abnormal nuclei of 0.67.

## I. Introduction

The chances of reverting a cancer case are strongly related to how early it is detected and treated [1]. Considering oral mucosa tumors, the early diagnosis can increase survival rates from 19% to 80% [2]. This disease is a global health problem, with over 350,000 people being affected each year [3] and the high mortality rate is mostly related to the late diagnosis due to the fact this disease doesn’t present symptoms at the early stages of development. Nonetheless, signs are present at the oral mucosa since the early beginning of the disease development and the use of non-invasive methods such as exfoliative exams are strongly recommended.

Currently, the biopsy is recognized as the gold-standard method for oral cancer diagnosis, but it is an invasive surgical procedure that is not recommended to be used as a screening test. On the other hand, the cytology exam is cheaper and doesn’t require special infrastructure because it is a non-invasive method for cell collection. Cytology is widely used as a diagnostic tool in medicine and dentistry and requires a stain for cells visualization (the most popular is the Papanicolaou Staining method), a light microscope for image amplification, and a Pathology consultant.

The early stages of cancer development can be identified by morphometric alterations in the affected cells’ nucleus. Given that these alterations seem to have an important role in cancer development, the morphological analysis using Papanicolaou is a promising and low-cost method for the early detection of malignant tumors. Besides that, this technique is not widely used in the clinical odontological routine because of the lack of an automated method for the analysis of this type of data [2]. Training the professionals that analyze these exams takes years of practice. Diagnose on cytology samples is still considered a manual and time-consuming process, subject to variations in perceptions and human error. In order to avoid these problems, computer-assisted analysis such as deep learning methods can be used to detect alterations of cellular status, helping on early cancer diagnosis [4].

### A. Objectives

Machine Learning has been employed to develop methods for the automated support of quantitative analysis of cytological samples for more than 25 years [5]–[9]. However, recent advances in the field of Deep Learning, have opened many new possibilities for the automated analysis of this type of image.

Segmentation, detection and classification approaches have already been applied with Papanicolaou [10], AgNOR [11] and Feulgen [12] slide images. In this work, our main objective is to evaluate the detection approach applied to a larger dataset of Papanicolaou stained oral cytology slide images. For this, we trained and compared, in terms of precision and recall, 4 deep learning detection models to evaluate the performance of this method in the cytology images domain.

## II. Related Works

The use of computer-aided technology in the analysis of oral cytology exams is not a well-explored area. To identify the state-of-the-art in this field, we analyzed two distinct Systematic Literature Reviews: [13], published in December 2019 as a technical report, and [14], published in May 2021. These reviews are focused on methods used to detect anomalies in cytology exams in oral examinations and other body parts.

In [14], the authors identified a growing tendency in researches using Deep Learning in the digital cytology field. Besides that, classic computer vision methods are still widely used. We found only two public datasets of Papanicolaou stained images using the conventional sample preparation technique, with only one containing slide fields with multiple cells and none with oral mucosa exams. Also, the analyzed works are in an experimental phase, without experiments in the clinical routine. Considering the automated analysis of oral cytology, in [13] the authors concluded that the research is mainly based on generic feature extraction methods as histogram analysis, active contours, and grayscale co-occurrence matrices. Those features are then used along with classification algorithms like ANN, SVM, and k-NN and no state-of-the-art methods like Deep Learning were applied to oral cytology slides images.

In [10] the authors compared models for three deep learning approaches: segmentation, object detection, and image classification. The results show that the binary object detection with Faster R-CNN was the best approach for nuclei detection and localization (0.76 IoU). Since ResNet 34 had a good performance on abnormal nuclei classification (0.88 accuracy, 0.86 *F*_1_ score) the authors concluded that these two models can be used in combination to make a localization and classification pipeline. This work reinforced that the automated analysis of oral cytology to make a pipeline for nuclei classification and localization using deep learning can help to minimize the subjectivity of the human analysis and also to detect cancer at early stages. In [15] the authors evaluated the segmentation and detection approaches for cell nuclei localization using the second version of the UFSC OCPap dataset. This work reinforced the Faster R-CNN as the best model for localization, with 0.81 of IoU.

## III. Material and Methods

To evaluate possible methods to analyze Papanicolaou stained slides, we applied the object detection approach. Object detection aims to provide each object of interest a bounding box with a label and can localize and classify nuclei. The code used in this work is publicly available at https://codigos.ufsc.br/lapix/segmentation-detection-and-classification-of-cell-nuclei-on-oral-cytology-samples-stained-with-papanicolaou.

### A. Data

We used the UFSC OCPap (v3) dataset, available at https://arquivos.ufsc.br/d/5035aec3c24f421a95d0/, to train and validate the models. This dataset comprises 6,128 images of 1200×1600 pixels acquired from 4 slides of cancer diagnosed and 2 healthy oral brush samples stained with Papanicolaou and captured with an Axio Scan.Z1 microscope and a Hitachi HV-F202SCL camera. The slides were provided by the Hospital Dental Center of the University Hospital Professor Polydoro Ernani de São Thiago of Federal University of Santa Catarina (HU-UFSC) ^1^. Each slide originated an image of 214,000×161,000 pixels (0.111*µm* x 0.111*µm* per pixel) that we divided into the fields that originated the dataset.

A group of specialists labeled these images using the LabelBox [16] free software tool and we split it into three subsets: 70% for training (4,307 images), 15% for validation (902 images), and 15% for test (919 images). Those subsets were created to evaluate the generalization capability of the trained model and to avoid biased results. The only set used to train the models was the training subset. The validation set was used for hyperparameter tuning during training, influencing the learning rate and the weight decay values. In this case, the validation set also influences the training process, which can result in overfitting. To avoid that, a more robust approach is the utilization of a third distinct set for evaluation of the results: the test subset [17]. Also, around 30% of the labels were reviewed by an experienced pathologist to assure the quality of the dataset. Figure 1 shows an example of Ground Truth (GT) and the respective label mask from the dataset. This figure exemplifies the complexity and diversity of the objects of interest in this type of image.

**Fig. 1.**
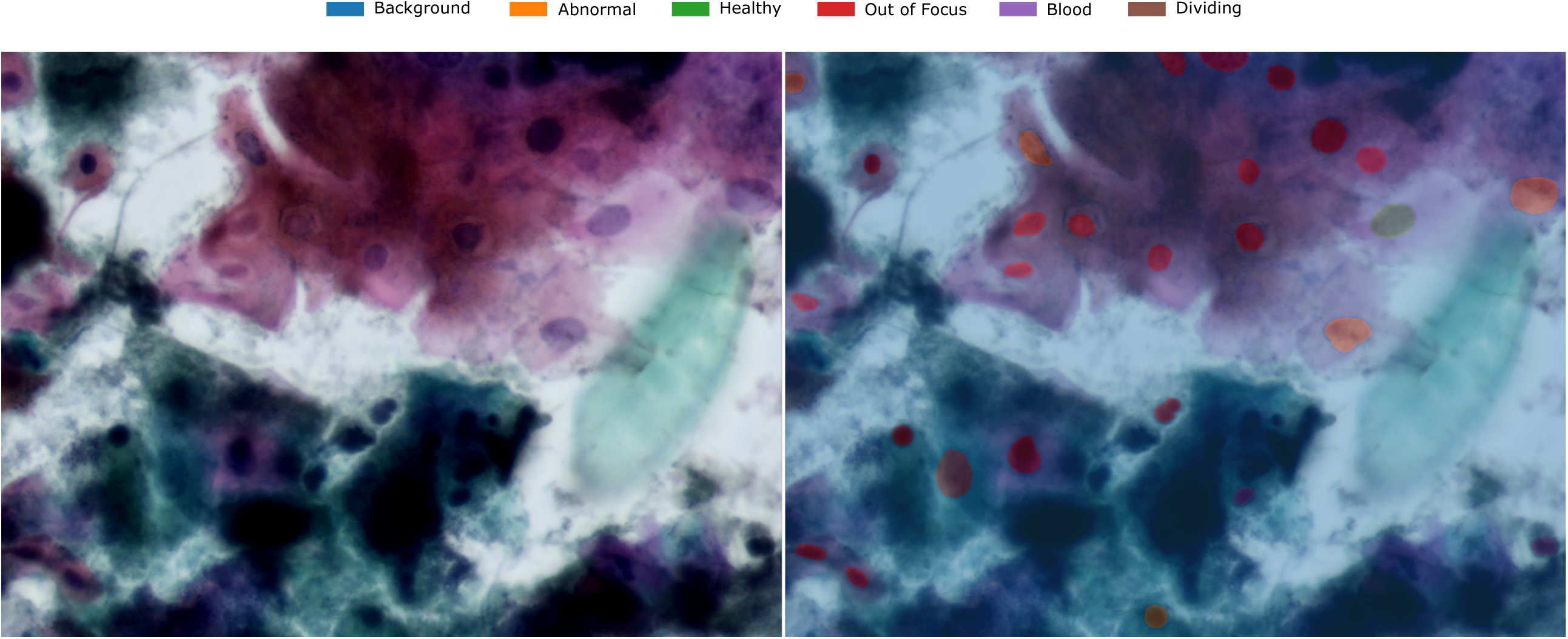
Example of Ground Truth (GT) and label mask from the UFSC OCPap (v3) dataset.

For this work, we used the entire dataset (*n = 6,128*). The labeling process generated bounding boxes labeled as “abnormal epithelial nucleus”, “healthy epithelial nucleus”, “out of focus nucleus”, “blood cell nucleus”, “reactive nucleus” and “dividing nucleus” for each image at each subset. Figure 2 shows the distribution of classes on each subset.

**Fig. 2.**
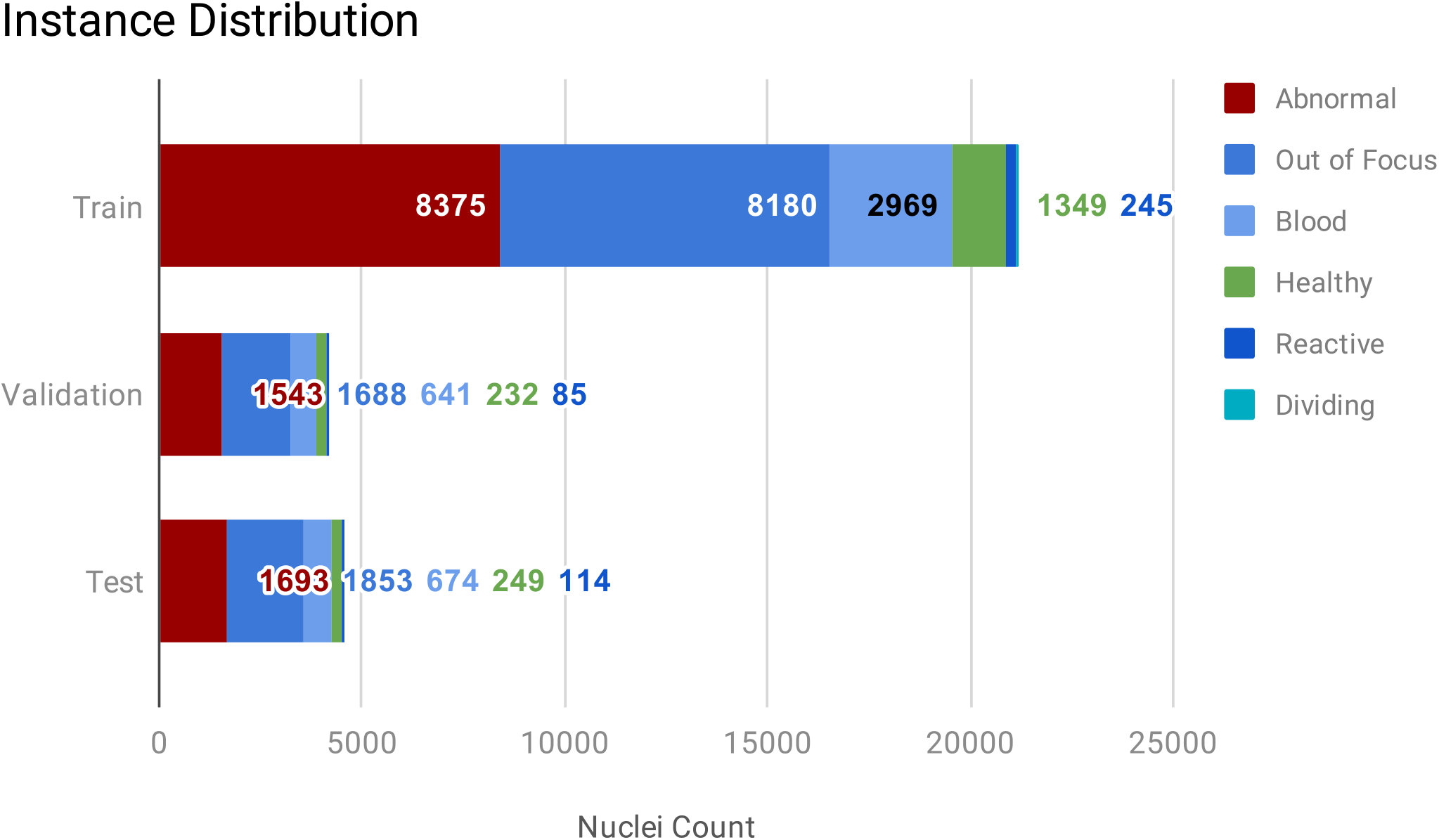
Chart of the nucleus instances distribution in the dataset.

### B. Experimental Setup

To train and validate the models, we employed the Detectron2 [18] API. We trained the networks using Nvidia Tesla K80 (12 GiB VRAM), T4 (16 GiB VRAM), and P100 (16 GiB VRAM) graphics cards, with an Intel(R) Xeon(R) CPU @ 2.20GHz × 4, 26 GiB of RAM and Ubuntu 18.04.3 LTS 64-bit. The models were pre-trained on COCO Dataset [19]. We trained 4 object detection models: Faster R-CNN [20], Mask R-CNN [21] and RetinaNet [22] with ResNet 50 and FPN [23] as backbone as they have shown state-of-the-art performance at Object Detection tasks on various image modalities. We selected the learning rate and weight decay as 0.005 and 0.0001 evaluating multiple training runs and trained each model for 2000 iterations. We also used the smooth L1 loss function for bounding boxes and cross-entropy for class loss with an SGD optimizer and 2 images per batch. We applied the following data augmentations: resizing to 640×853, 672×896, 704×939, 736×976, 768×1024, and 800×1067, and random horizontal flip with a probability of 0.5.

## IV. Results and Discussion

To evaluate the models, the precision and recall were calculated for each individual class to better show the classification performance of the models. To evaluate the localization performance, the IoU was calculated for the bounding boxes disconsidering the classes along with the instance count. All metrics were calculated on the test set with an IoU threshold of 0.5 and a score threshold of 0.5. An example of the results of two images of the test set can be seen in Figure 3.

**Fig. 3.**
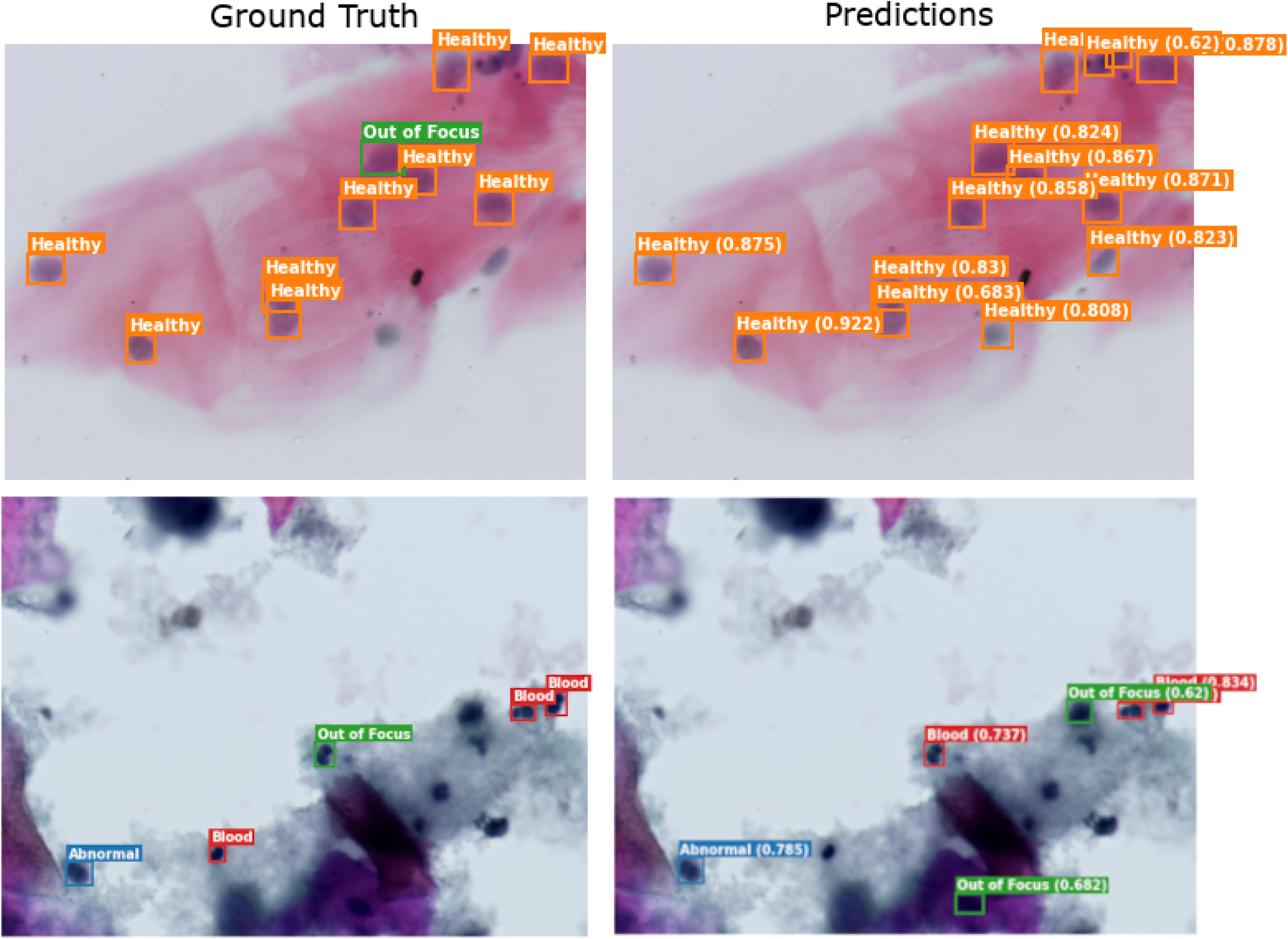
Predictions using Mask R-CNN on images of the test set.

Table I shows the precision and recall metrics of the models’ detections for each class. These results show that Mask R-CNN has the best recall for abnormal and normal nuclei. Considering that our objective is to detect abnormal cells, the abnormal nuclei recall is considered the most important metric in this work. For this reason, we considered that RetinaNet, despite having the best results for precision, had the worse results as it showed a low recall for both the main classes. Also, none of the models could detect reactive and dividing nuclei due to their under-representativity in the dataset.

**TABLE I.** Metrics results for all models by class. (P = Precision, R = Recall)

| Model | Abnormal |  | Healthy |  | Blood |  | Out of Focus |  |
| --- | --- | --- | --- | --- | --- | --- | --- | --- |
|  | P | R | P | R | P | R | P | R |
| Faster R-CNN | 0.59 | 0.46 | 0.72 | 0.56 | 0.43 | **0.53** | 0.34 | **0.42** |
| Mask R-CNN | 0.52 | **0.67** | 0.54 | **0.74** | **0.52** | 0.34 | 0.27 | 0.33 |
| RetinaNet | **0.68** | 0.40 | **0.83** | 0.34 | 0.46 | 0.12 | 0.48 | 0.01 |
| YOLO v4 CSP | **0.68** | 0.37 | 0.60 | 0.68 | 0.48 | 0.07 | **0.77** | 0.02 |

Figure 4 shows the localization IoU performance of all the evaluated models compared to the previous work that uses the second version of the dataset (v2). These results show that with the inclusion of more images, the IoU performance of the models became worse, even though Faster R-CNN is still better than RetinaNet for localization. Considering this metric, the Mask R-CNN model had betters results than Faster R-CNN, being the best model considering these metrics.

**Fig. 4.**
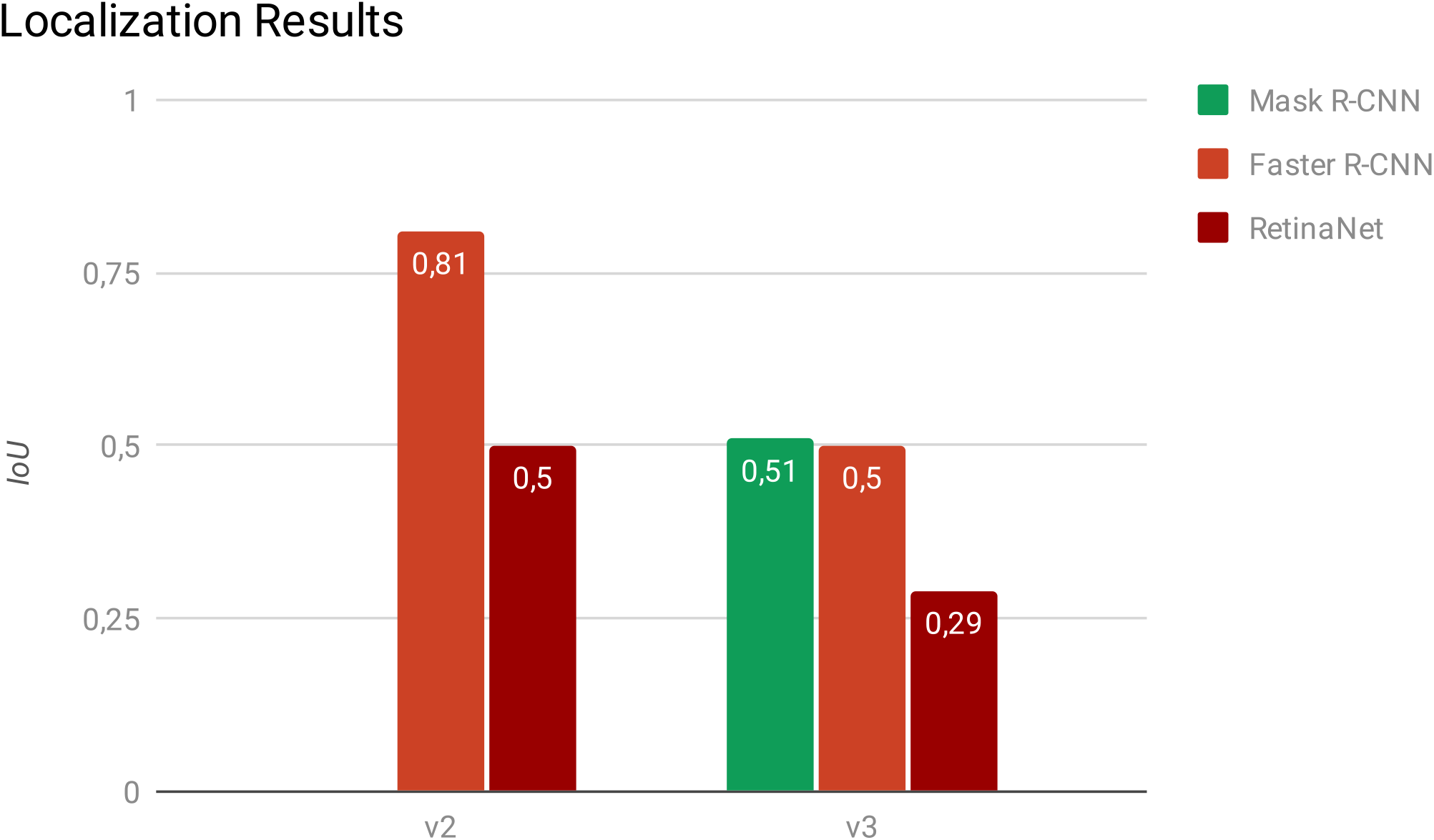
IoU detection results metrics.

Figure 5 provides instance-level statistics in comparison with the previous work (v2) along with the Mean Squared Error (MSE) per image of each model. The number of instances in the Ground Truth (GT) of the test set is represented by the green area. It is possible to see that Faster R-CNN had the best localization result, improving from the previous work results, followed by Mask R-CNN. This indicates that the models can localize the nuclei, despite having a low recall and precision for classification of the nuclei.

**Fig. 5.**
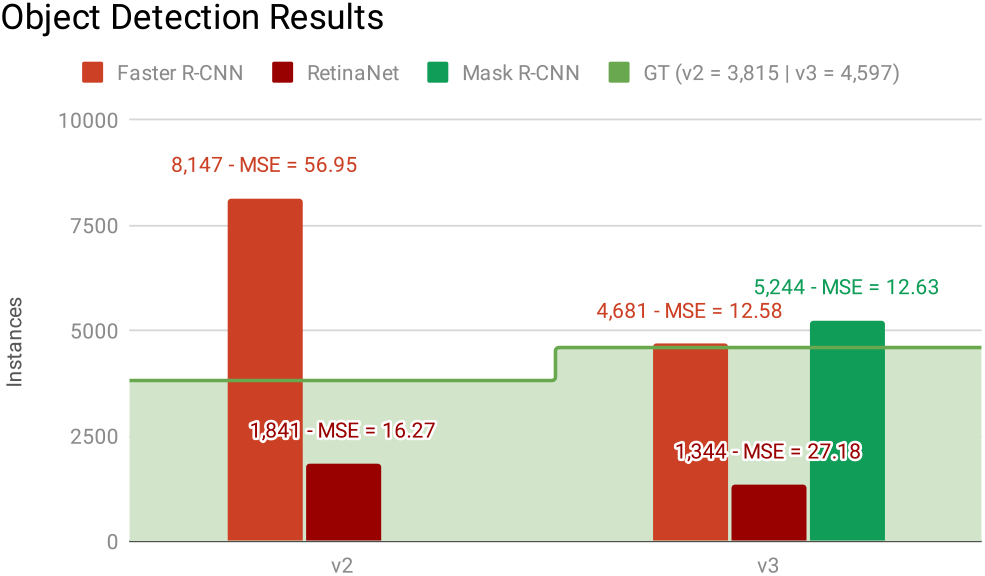
Instance level results chart.

Table II shows the results for abnormal cells of the related works extracted from [14] that use similar evaluation metrics to the ones that we employed for the evaluation of our models, and can be objectively compared to our results, despite using distinct datasets. Except for [24], all the cited related works use liquid-based samples. This sample preparation technique produces slide images with fewer overlapping cells and fewer artifacts, which are less complex to be analyzed by a computer vision method [24]. However, using this method has a higher cost when compared to the Conventional Pap Smear technique, used in our work, making it not viable in the public health systems of developing countries [25]. Considering that Liquid Based Cytology (LBC) Pap samples and Conventional Pap samples produce considerably different images, the papers are divided into these two groups. The best results for the IoU metric achieved 0.90 in LBC Pap smear slides, which emphasizes that it is easier to detect nuclei in this kind of samples while our work achieved the maximum 0.51 IoU using Conventional Pap Smear samples. Analyzing the *F*_1_ Score, our work had a similar result (0.51) with the work that had the best performance [24] (0.53) considering this metric for Conventional Pap Smear samples. It is also important to notice that two papers [26], [27] indicate that Faster R-CNN has the best results for images of Papanicolaou stained samples, both LBC and conventional, which corroborates with our results.

**TABLE II.**
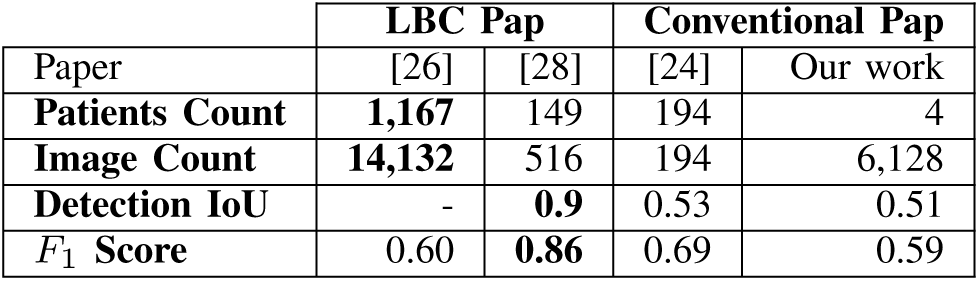
Best results obtained by the main related works for abnormal cells. The best results of each metric are highlighted in bold.

| Paper | LBC Pap |  | Conventional Pap |  |
| --- | --- | --- | --- | --- |
|  | [26] | [28] | [24] | Our work |
| **Patients Count** | **1,167** | 149 | 194 | 4 |
| **Image Count** | **14,132** | 516 | 194 | 6,128 |
| **Detection IoU** | - | **0.9** | 0.53 | 0.51 |
| **$F_1$ Score** | 0.60 | **0.86** | 0.69 | 0.59 |

### A. What is new here in relation to the previous works?

In comparison with the previous works, in this paper we applied one new detection model (Mask R-CNN) using a larger, more diverse, and more balanced version of the dataset, including 2 new slides of healthy cases and another 1 of a cancer case.

## V. Conclusion

In this work, we evaluated deep learning models for the object detection approach using the Faster R-CNN, Mask R-CNN, and RetinaNet models. Comparing the IoU, precision, and recall results, we concluded that the Mask R-CNN was the best model for localization and classification of the nuclei with an IoU of 0.51 and recall of abnormal nuclei of 0.67. Besides those results are not considered good enough for the automated detection of abnormal nuclei in Papanicolaou stained cytology, the two best models showed the capacity to localize the nuclei. These results reinforce what we concluded in the previous works: there seems to be necessary a composed pipeline, using separate models for localization and classification of the cells.

Hence, as future works, we plan to test an additional set of different algorithms, approaches, and methods to build a complete pipeline of automated slide analysis to help health professionals to early diagnose oral cancer cases. Furthermore, we want to enhance the quality and size of the dataset and generalize the analysis to other tissues and types of cell nuclei to support the diagnosis of other types of cancer. The objective is, in the next years, to integrate this pipeline into the Telepathology module [29] and the Oral Telemedicine module [30] of a public statewide Telemedicine system [31], [32] and to perform a large-scale validation study of our approach that will also consider the feasibility of integrating Tele-Cytology into the Telemedicine workflow.

The main contribution of this work is to show the performance of a possible solution for the automation of the analysis of oral cytology Papanicolaou stained slides using deep learning, highlighting the fact that the Papanicolaou staining method is also a promising approach for oral cancer diagnosis. We believe that our research can help to spread the Papanicolaou method as a screening exam to detect oral cancer at early stages and improve the treatment success.

## Data Availability

We used the UFSC OCPap (v3) dataset, available at https://arquivos.ufsc.br/d/5035aec3c24f421a95d0/, to train and validate the models.

## Acknowledgements

We want to thank Fundação de Amparo à Pesquisa e Inovação do Estado de Santa Catarina (Fapesc) and Coordenação de Aperfeiçoamento de Pessoal de Nível Superior (CAPES) for funding this work. Also, we want also to thank Prof. Felipe Perozzo Daltoé for providing the samples and to review the labels and Bruna Zanicoski, Filipe Landa, Ricardo Pereira, and Ricardo Thisted for labeling the images.

## Footnotes

1 This research was approved by the UFSC Research Ethics Committee (CEPSH), protocol number 23193719.5.0000.0121. All patients were previously approached and informed about the study objectives. Those who agreed to participate signed an Informed Consent Form.

